# The TRIPOD-LLM Statement: A Targeted Guideline For Reporting Large Language Models Use

**DOI:** 10.1101/2024.07.24.24310930

**Authors:** Jack Gallifant, Majid Afshar, Saleem Ameen, Yindalon Aphinyanaphongs, Shan Chen, Giovanni Cacciamani, Dina Demner-Fushman, Dmitriy Dligach, Roxana Daneshjou, Chrystinne Fernandes, Lasse Hyldig Hansen, Adam Landman, Lisa Lehmann, Liam G. McCoy, Timothy Miller, Amy Moreno, Nikolaj Munch, David Restrepo, Guergana Savova, Renato Umeton, Judy Wawira Gichoya, Gary S. Collins, Karel G. M. Moons, Leo A. Celi, Danielle S. Bitterman

## Abstract

Large Language Models (LLMs) are rapidly being adopted in healthcare, necessitating standardized reporting guidelines. We present TRIPOD-LLM, an extension of the TRIPOD+AI statement, addressing the unique challenges of LLMs in biomedical applications. TRIPOD-LLM provides a comprehensive checklist of 19 main items and 50 subitems, covering key aspects from title to discussion. The guidelines introduce a modular format accommodating various LLM research designs and tasks, with 14 main items and 32 subitems applicable across all categories. Developed through an expedited Delphi process and expert consensus, TRIPOD-LLM emphasizes transparency, human oversight, and task-specific performance reporting. We also introduce an interactive website (https://tripod-llm.vercel.app/) facilitating easy guideline completion and PDF generation for submission. As a living document, TRIPOD-LLM will evolve with the field, aiming to enhance the quality, reproducibility, and clinical applicability of LLM research in healthcare through comprehensive reporting.

**COI:** DSB: Editorial, unrelated to this work: Associate Editor of Radiation Oncology, HemOnc.org (no financial compensation); Research funding, unrelated to this work: American Association for Cancer Research; Advisory and consulting, unrelated to this work: MercurialAI. DDF: Editorial, unrelated to this work: Associate Editor of JAMIA, Editorial Board of Scientific Data, Nature; Funding, unrelated to this work: the intramural research program at the U.S. National Library of Medicine, National Institutes of Health. JWG: Editorial, unrelated to this work: Editorial Board of Radiology: Artificial Intelligence, British Journal of Radiology AI journal and NEJM AI. All other authors declare no conflicts of interest.

## Introduction

Healthcare’s embrace of Large Language Models (LLMs) shows no signs of slowing down, with current and future deployment being considered in several domains across administrative and healthcare delivery use-cases, including in-basket draft generation, medical document summarization, question answering, information retrieval, medical diagnosis, treatment recommendations, patient education, and medical education.^1–5^ The rapid advancements made in LLMs have stretched existing regulatory and governance structures to their limits, exposing a patchwork of solutions that do not fully encompass the nuances of these all-purpose models.^6–8^ More broadly, with this speed, LLMs have posed a challenge to journal and peer-review publication timelines and challenged regulatory agencies to provide timely guidance. To maintain the speed, researchers publish pre-prints quickly and take an ad-hoc approach to reporting.

Reporting guidelines provide a scalable method for standardizing research, transparent reporting, and the peer review process. The TRIPOD (Transparent Reporting of a Multivariable Model for Individual Prognosis Or Diagnosis) initiative is a critical example that was first introduced in 2015 to establish minimum reporting standards for diagnostic and prognostic prediction model studies (www.tripod-statement.org).^9^ TRIPOD is one of the core guidelines on the EQUATOR Network, which is an international effort that promotes transparent, accurate reporting of health research literature.^10^ TRIPOD is widely endorsed and recommended by journals, and is often included in journal instructions to authors. TRIPOD has subsequently been updated to incorporate best practices for AI due to the significantly evolved machine learning landscape, resulting in TRIPOD+AI.^11^ This is in addition to other guidelines that offer complementary guidance on AI development throughout the model lifecycle.^12–14^ However, LLMs represent a distinct frontier within AI, introducing unique challenges and considerations not fully addressed by original TRIPOD guidelines or its newer extensions as we shift from classifier AI models to generative AI. Here, we report the TRIPOD+LLM statement, an extension of TRIPOD+AI^11^, developed to address these unmet needs and designed to be a living checklist in order to nimbly adapt to the rapidly evolving field. A completed example from a recent LLM research study is provided to guide future users (Supplementary Table 1).

### LLMs for biomedicine introduce unique complexities

LLMs as generative AI models are autoregressive, meaning they are trained to predict the next word in a sequence given the words that preceded it. Yet, this foundational training has been shown to equip them with capabilities to perform a wide range of healthcare-related natural language processing (NLP) tasks from a single model. This adaptability is commonly achieved through supervised fine-tuning (SFT) or few-shot learning methods, which allow LLMs to handle new tasks with minimal examples.^15,16^ Chatbot solutions (e.g., ChatGPT) use LLMs as their foundation, upon which two more components are added: question-answering (referred to as instruction tuning or supervised fine-tuning) and preference ranking (referred to as alignment). The unique methodological processes involved in LLMs and chatbots are not captured by current guidelines, such as the choice of hyperparameters used for SFT, the intricacies of prompting, variability in model predictions, methods in evaluating natural language outputs, and preference-based learning strategies, which require specific guidance and significantly impact model reliability. In addition, the generalist and generative nature of LLMs require more detailed guidance than covered in prior guidelines. Because LLMs can be applied to a broad range of use cases for which they were not specifically trained for and were not necessarily represented in training data (e.g. disease prevalence typically captured in a task-specific model’s training data for a given use case), they require unique task-specific guidance for robust reporting and downstream reliability and safety.

The selection of appropriate automated and human metrics by which to evaluate generative output remains an open question, and currently a wide range of methodologies are applied to capture various facets of performance. For tasks where the output is truly unstructured text and cannot be resolved to a structured label, evaluation is particularly complex. In these cases, most automated metrics prioritize overlap and similarity between input and output text, producing scores that may not accurately capture factual accuracy or relevance of the text produced, especially hallucinations or omissions.^17–19^ These scores reflect the degree of structural and lexicographical resemblance which, though important, capture only a fraction of what constitutes a comprehensive evaluation of performance and safety. Human evaluation of text is subject to subjective interpretation, complicated by the ambiguity of language and uncertainties inherent to many clinical tasks. These challenges are heightened in medicine, where there is often no single correct answer and both aleatoric and epistemic uncertainty are common. Therefore, more details are needed to guide reporting of how performance is evaluated. In this paper, we use the term LLM to refer to both LLMs and chatbots. Table 1 highlights key categories of tasks applicable to the healthcare domain and provides notable definitions and examples of existing relevant work.

**Table 1.**
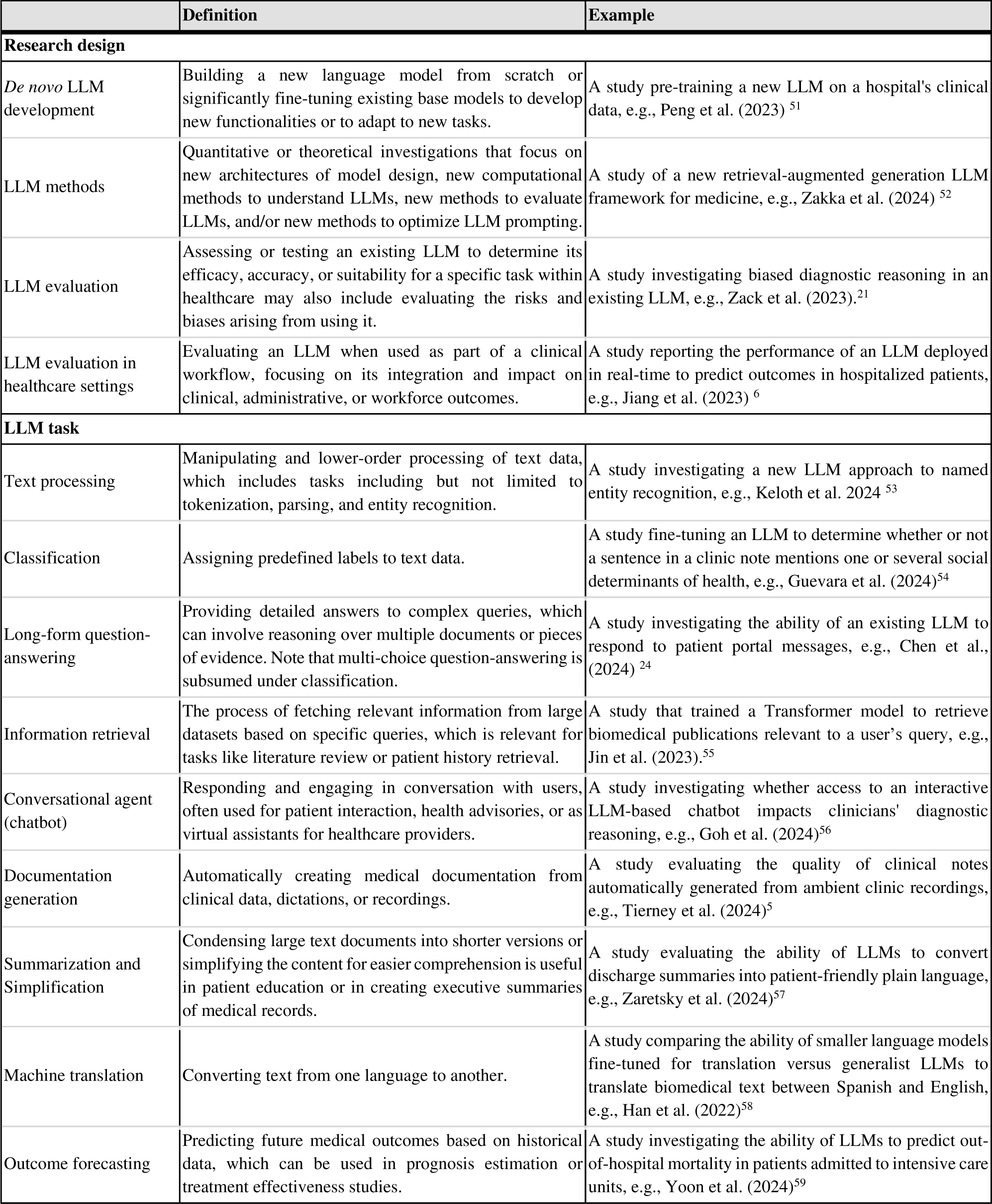
Research design and LLM task categories for the modular TRIPOD-LLM guideline.

The novel complexities introduced by LLMs include concerns regarding hallucinations, omissions, reliability, explainability, reproducibility, privacy, and biases being propagated downstream, which can adversely affect clinical decision-making and patient care.^20–26^ Furthermore, growing partnerships between EHR vendors, technology companies, and healthcare providers have led to deployment horizons that far outpace current regulatory timelines.^8,27^ To safeguard LLM use and increase transparency, standardization in developing and reporting LLMs is essential to ensure consistency, reliability, and verifiability, akin to established clinical grade evaluation in other scientific domains.^28–30^

## Results

### The TRIPOD-LLM Statement

The TRIPOD-LLM comprises a checklist of items considered essential for good reporting of studies that are developing, tuning, prompt engineering, or evaluating an LLM (Table 2). Box 2 summarizes noteworthy additions and changes to TRIPOD-2015 and TRIPOD+AI. The TRIPOD-LLM checklist comprises 19 main items about the title (1 item), abstract (1 item), introduction (2 items), methods (8 items), open science practices (1 item), patient and public involvement (1 item), results (3 items), and discussion (2 items). These main items are further divided into 50 subitems. Of these, 14 main items and 32 subitems are applicable to all research designs and LLM tasks. The remaining 5 main items and 18 subitems are specific to particular research designs or LLM task categories. As discussed in the methods, the TRIPOD-LLM statement introduces a modular format given the varied nature of LLM studies (Table 1), where some items are only relevant for specific research designs and LLM task categories.

**Table 2:**
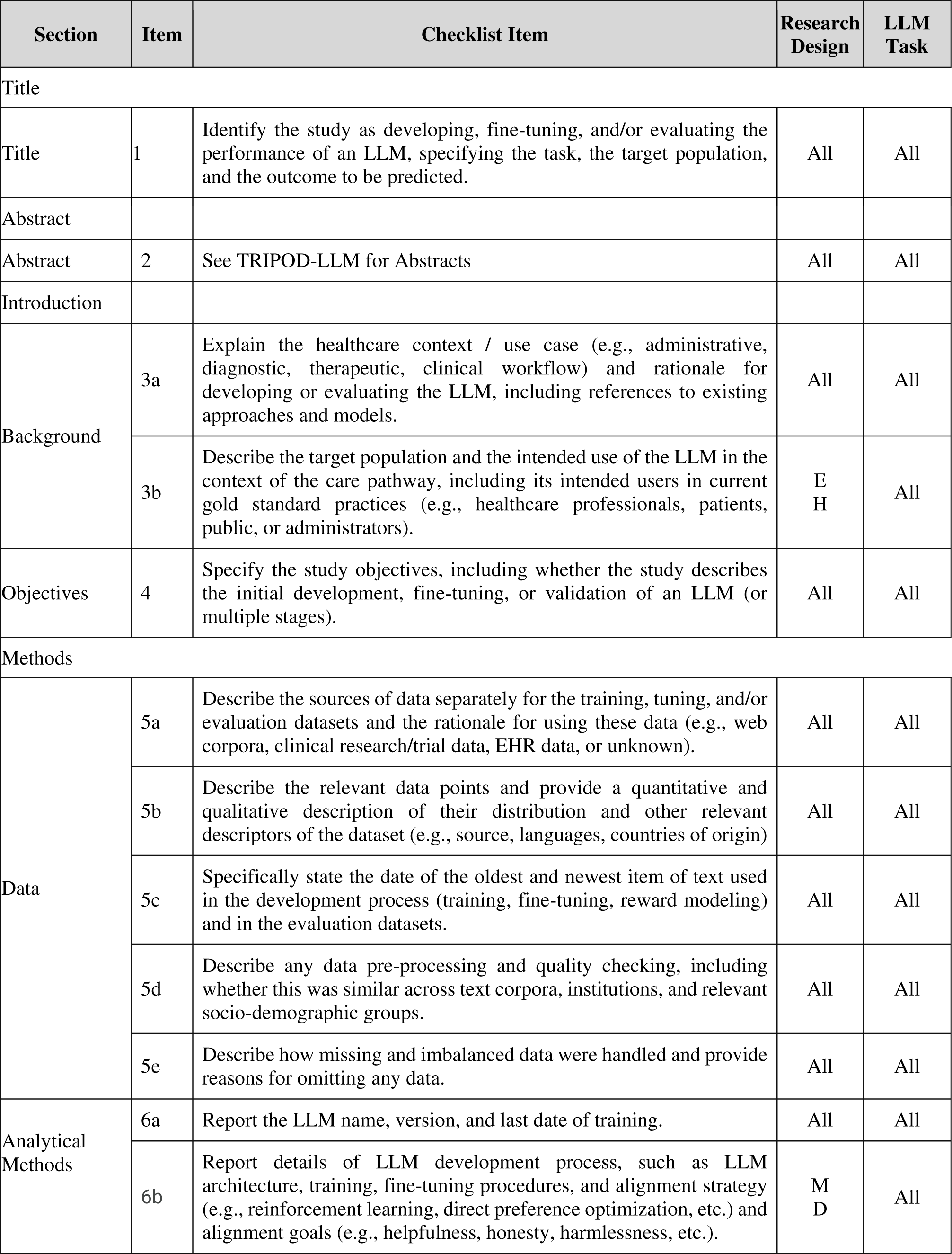
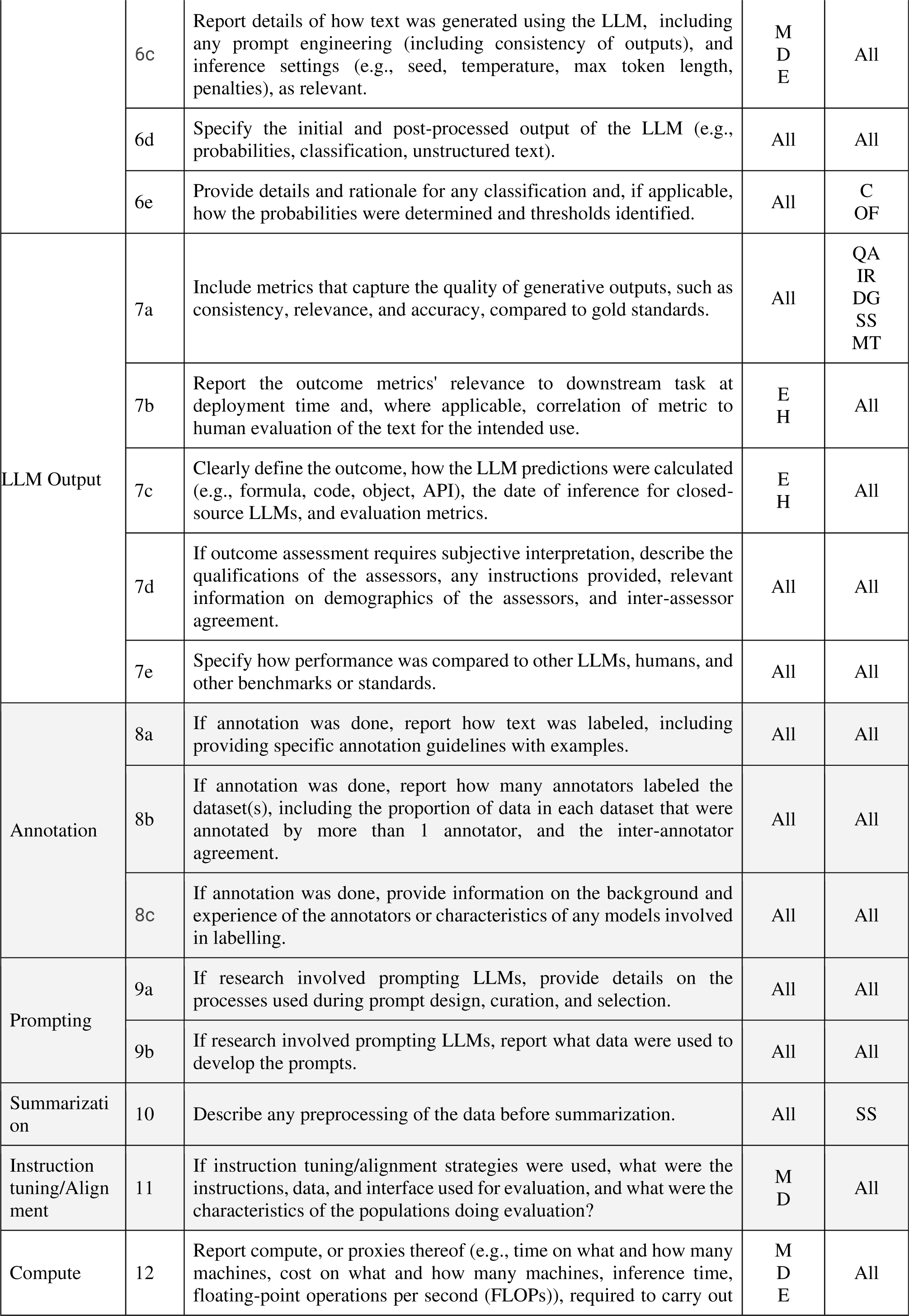
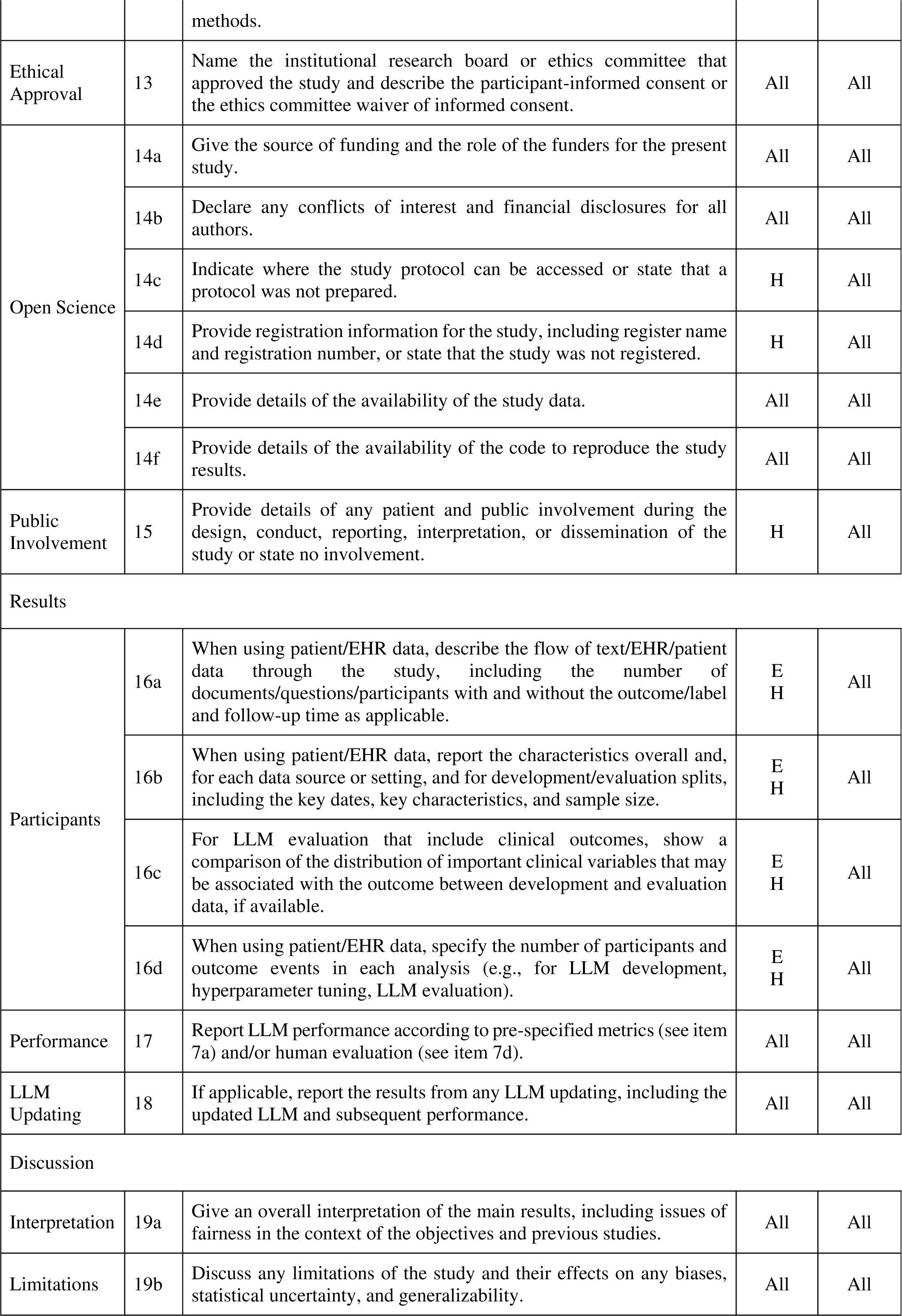
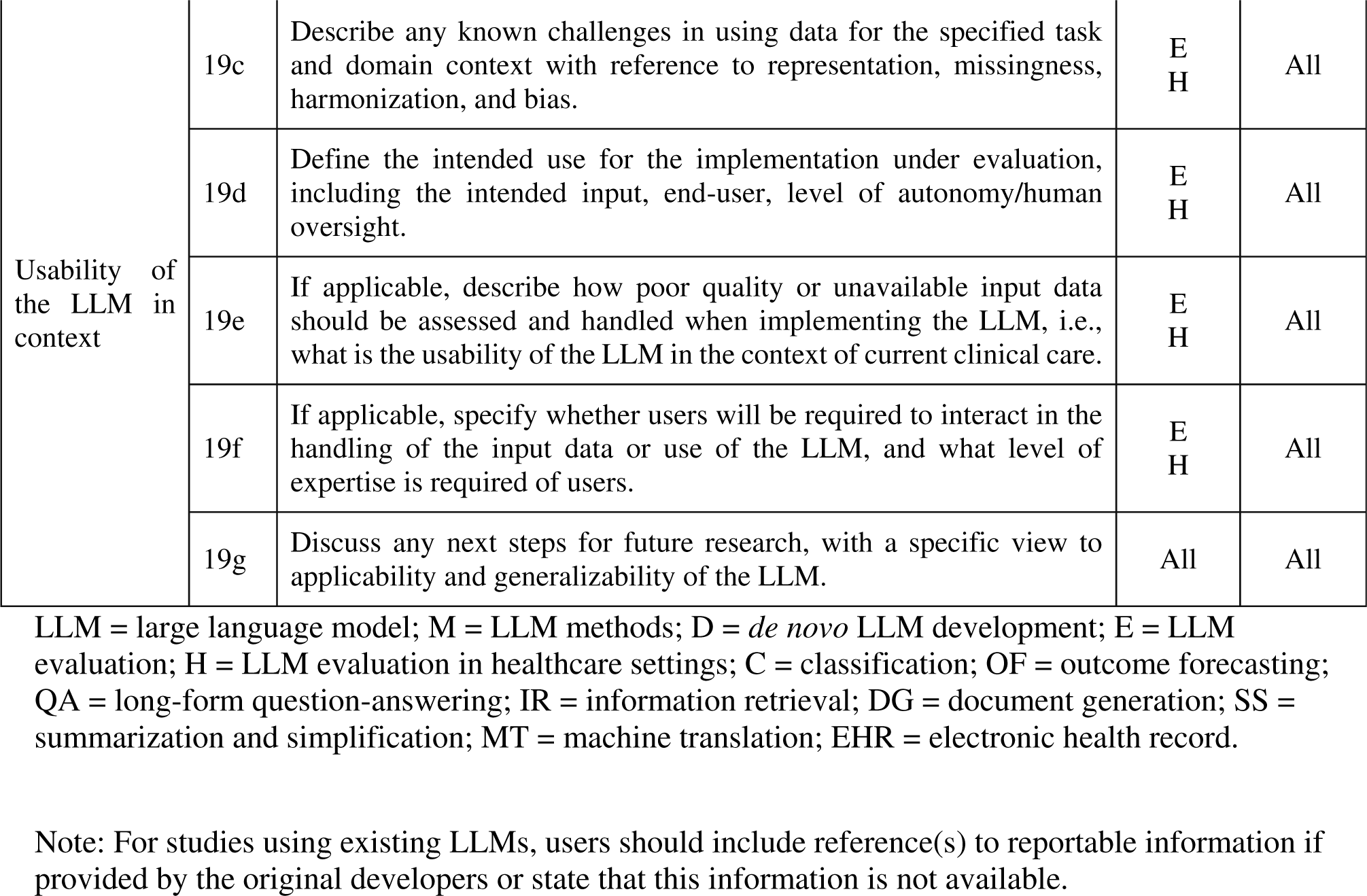
TRIPOD-LLM Checklist.

#### Box 1: Glossary of terms (in alphabetical order)

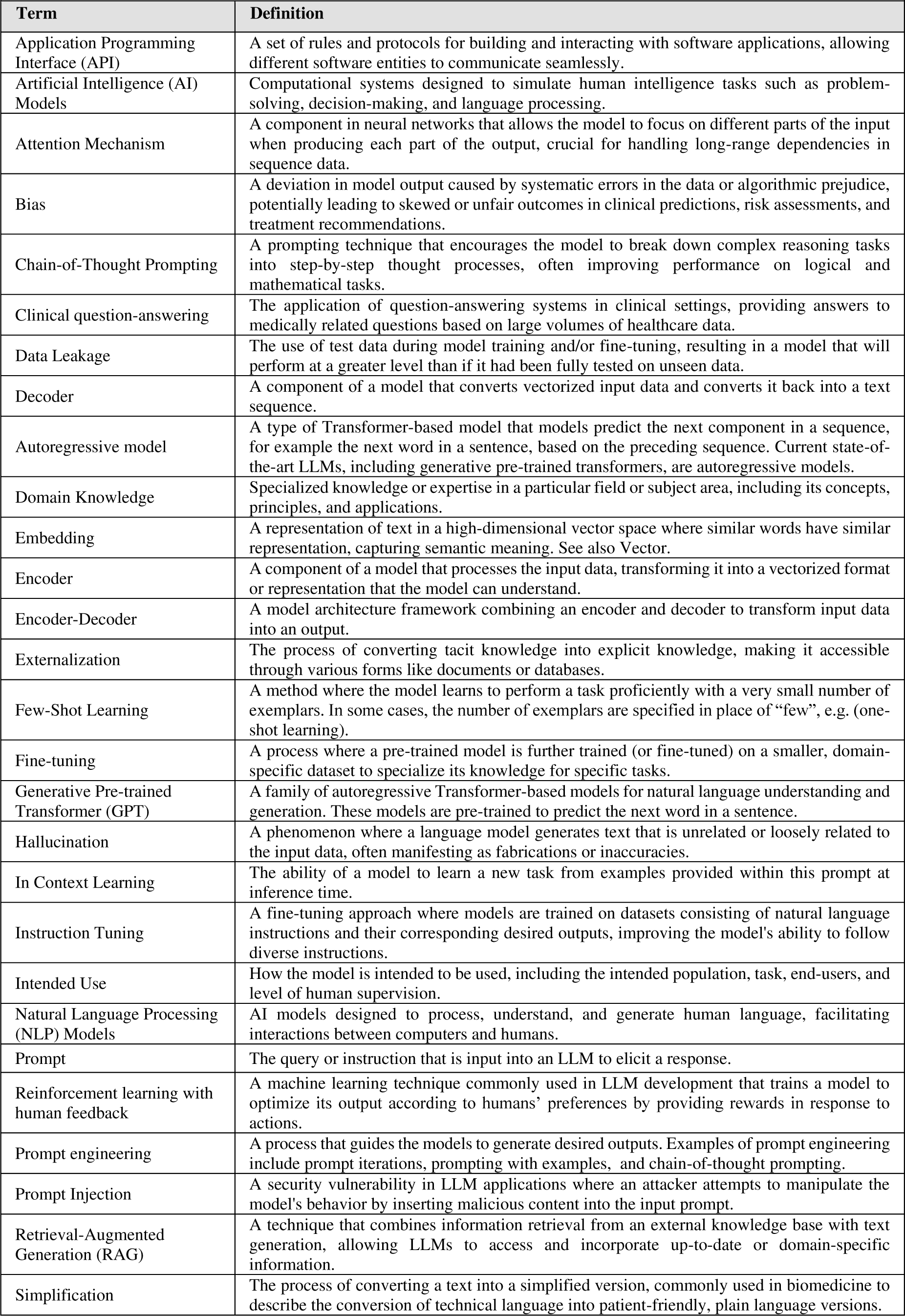
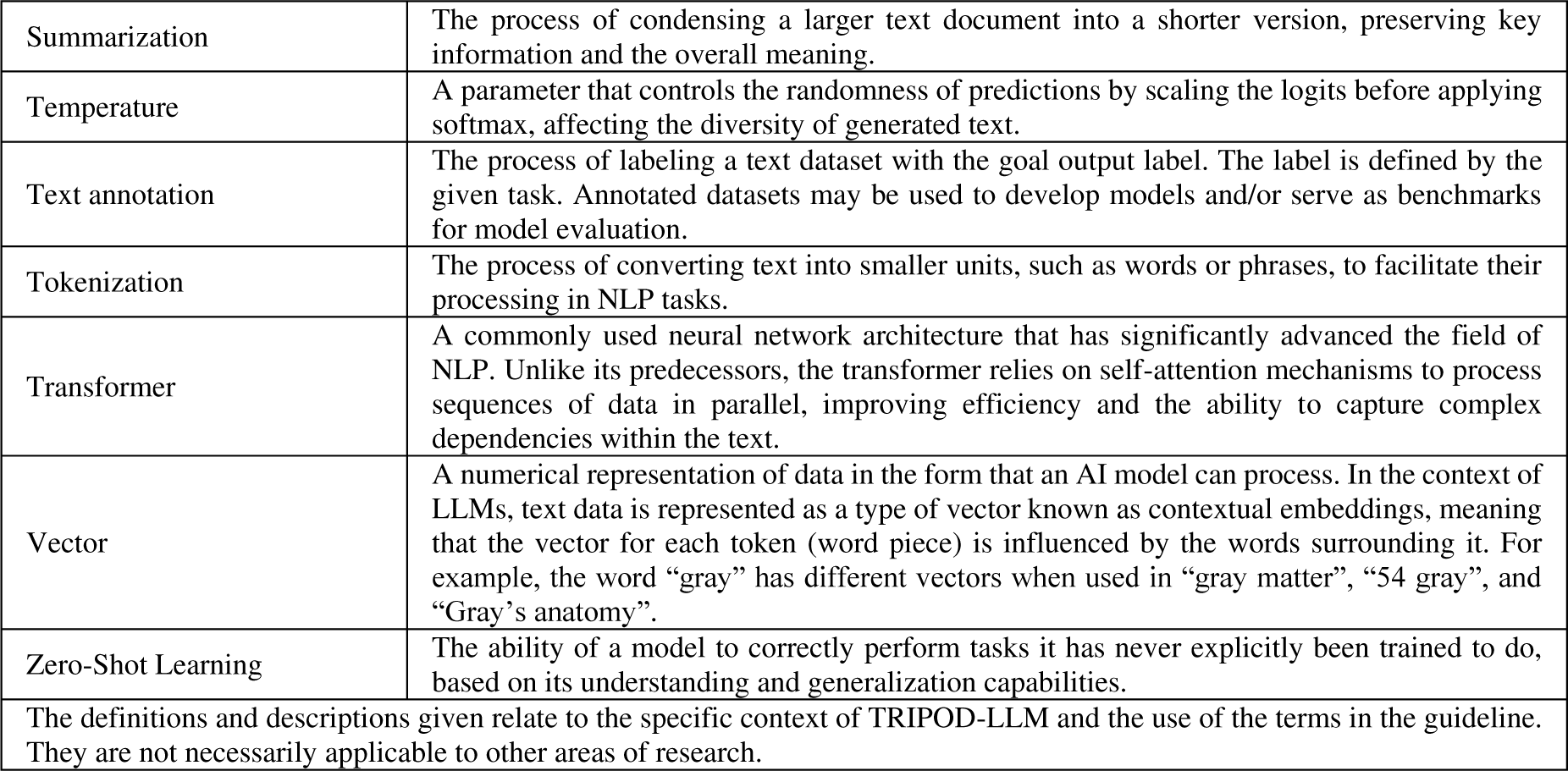

#### Box 2: TRIPOD-LLM noteworthy changes and additions to TRIPOD-2015 and TRIPOD+AI

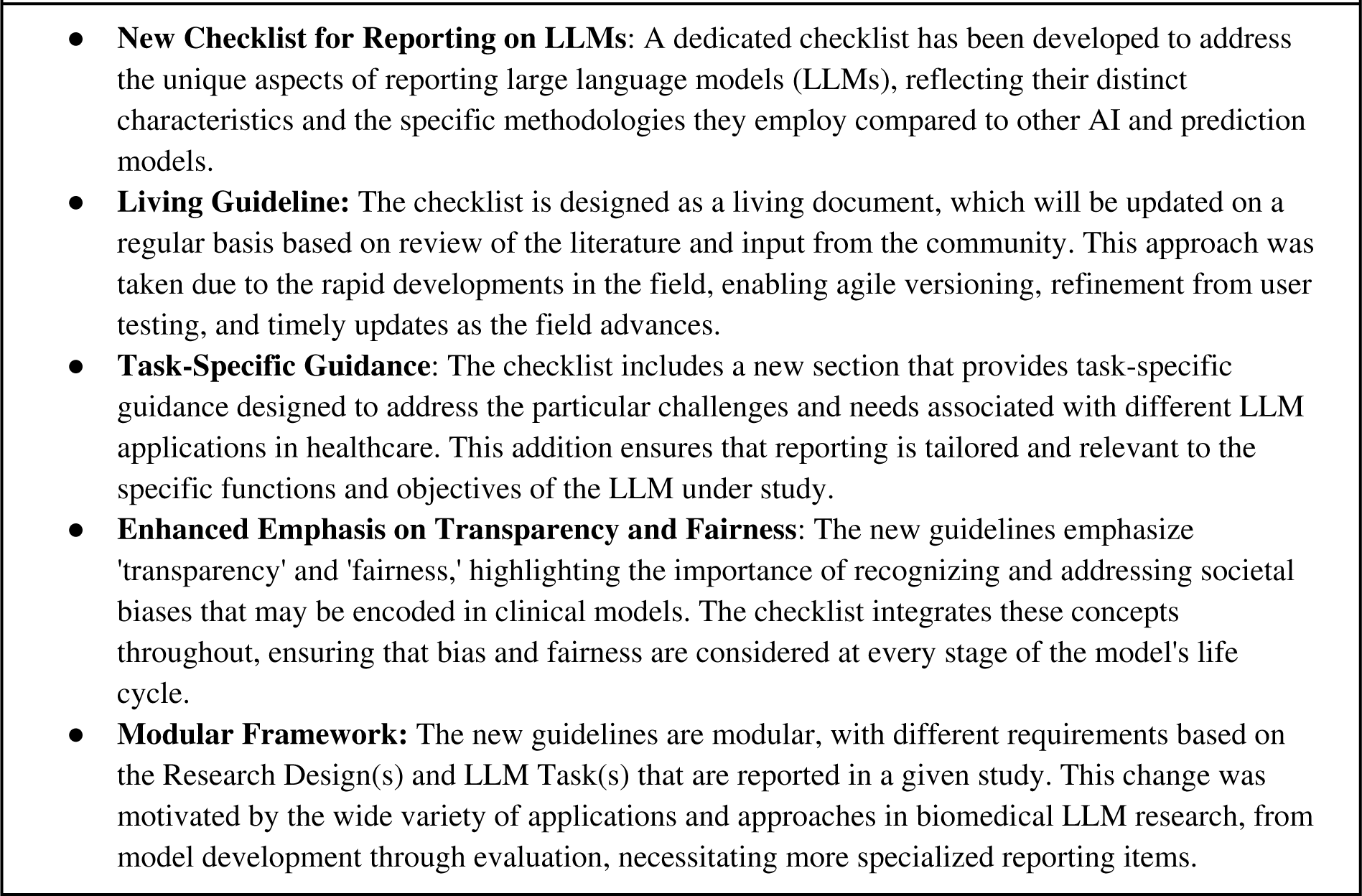

A separate checklist for journal or conference abstracts of LLM-based studies is included, and the TRIPOD+AI for Abstracts statement^18^ is revised (TRIPOD-LLM for Abstracts), reflecting new content and maintaining consistency with TRIPOD-LLM (Table 3).

**Table 3:**
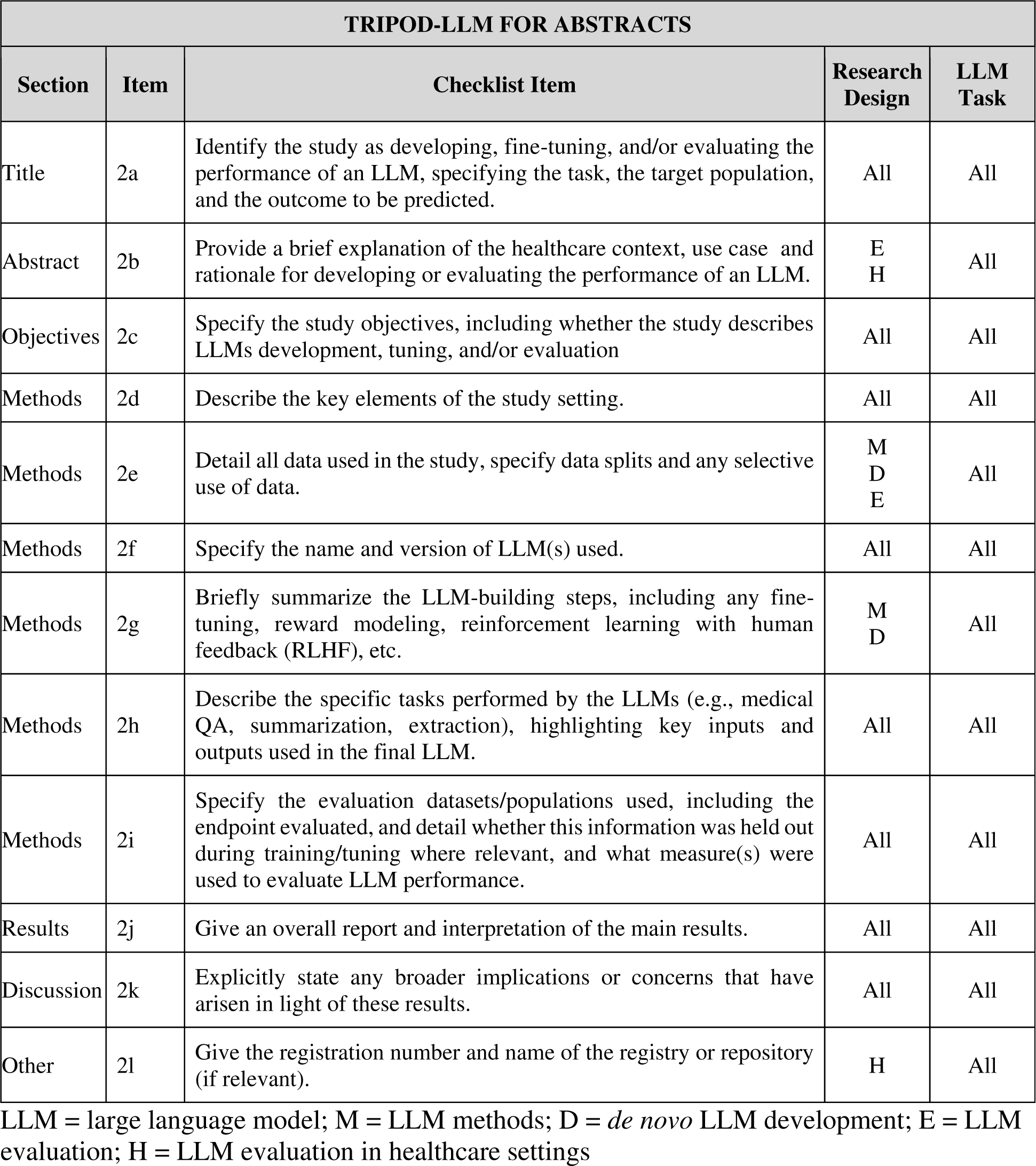
Essential items to include in a journal or conference abstract for a study describing the development, fine-tuning, or evaluation of a large language model (TRIPOD-LLM for Abstracts*)

| TRIPOD-LLM FOR ABSTRACTS |  |  |  |  |
| --- | --- | --- | --- | --- |
| Section | Item | Checklist Item | Research Design | LLM Task |
| Title | 2a | Identify the study as developing, fine-tuning, and/or evaluating the performance of an LLM, specifying the task, the target population, and the outcome to be predicted. | All | All |
| Abstract | 2b | Provide a brief explanation of the healthcare context, use case and rationale for developing or evaluating the performance of an LLM. | E<br>H | All |
| Objectives | 2c | Specify the study objectives, including whether the study describes LLMs development, tuning, and/or evaluation | All | All |
| Methods | 2d | Describe the key elements of the study setting. | All | All |
| Methods | 2e | Detail all data used in the study, specify data splits and any selective use of data. | M<br>D<br>E | All |
| Methods | 2f | Specify the name and version of LLM(s) used. | All | All |
| Methods | 2g | Briefly summarize the LLM-building steps, including any fine-tuning, reward modeling, reinforcement learning with human feedback (RLHF), etc. | M<br>D | All |
| Methods | 2h | Describe the specific tasks performed by the LLMs (e.g., medical QA, summarization, extraction), highlighting key inputs and outputs used in the final LLM. | All | All |
| Methods | 2i | Specify the evaluation datasets/populations used, including the endpoint evaluated, and detail whether this information was held out during training/tuning where relevant, and what measure(s) were used to evaluate LLM performance. | All | All |
| Results | 2j | Give an overall report and interpretation of the main results. | All | All |
| Discussion | 2k | Explicitly state any broader implications or concerns that have arisen in light of these results. | All | All |
| Other | 2l | Give the registration number and name of the registry or repository (if relevant). | H | All |

LLM = large language model; M = LLM methods; D = *de novo* LLM development; E = LLM evaluation; H = LLM evaluation in healthcare settings

The recommendations contained within TRIPOD-LLM are for completely and transparently reporting on how LLM-based research was conducted; TRIPOD-LLM does not prescribe how to develop or evaluate LLMs specifically. The checklist is not a quality appraisal tool. Similarly, CANGARU ^31^ and CHART ^32^ are complementary guidelines that relate to Generative AI more broadly and Chatbots specifcially.

In addition to the TRIPOD website (www.tripod-statement.org) an accompanying interactive website was developed (https://tripod-llm.vercel.app/) to present the required questions based on research design(s) and task(s) for ease of completion. This site can be used to render a final PDF suitable for submission. Fillable templates for the TRIPOD-LLM checklist can also be found in the supplementary material or downloaded from www.tripod-statement.org. News, announcements, and information relating to TRIPOD-LLM and the release of subsequent statements can be found on the TRIPOD-LLM website, TRIPOD website (www.tripod-statement.org), and on social media accounts such as X (formerly known as Twitter) @TRIPODStatement.

An example of a completed TRIPOD-LLM checklist for a previously published study reporting the pre-training, fine-tuning, retrospective evaluation, and clinical deployment of an LLM for clinical and operational hospital tasks is presented in Table 5. The design categories relevant to this work are *de novo* LLM development, LLM evaluation, and LLM evaluation in healthcare settings. Task categories relevant to this work are classification and outcome forecasting.

### TRIPOD-LLM Statement as a Living Document

Given the rapid pace of the field and the timeline for interaction with healthcare workers and patients, the decision was made to create an accelerated TRIPOD-LLM statement to provide timely guidance for LLM use in (bio)medical and other healthcare applications. This guidance has been designed as part of a living document hosted on an interactive website to facilitate agile versioning, refinement from user testing, updates as the field evolves, and regular meetings to intake and evaluate new standards. Thus, as the reporting recommendations are anticipated to evolve; users are directed to the most current version of the guidelines at https://tripod-llm.vercel.app/.

Our approach to the living TRIPOD-LLM statement is informed by processes established in developing living systematic reviews ^33,33,34^ and clinical practice guidelines,^35,36^ which have been adopted to address a similar need to provide updated, timely recommendations based on evolving evidence. Public comments on the statement will be collected from the community via multiple avenues to enhance accessibility: a project-specific GitHub repository, the TRIPOD-LLM website, and the main TRIPOD website (https://www.tripod-statement.org/). We encourage input both on usability, such as language that may be ambiguous or redundant, and on the content of the guidelines themselves. As a few examples, users may suggest a change to an item to make it more feasible in practice, recommend a new item be added, recommend adding or removing items assigned to a given research design of LLM task module, or recommend changes to the research design or LLM task module categories.

An expert panel will convene every three months to discuss updates. Before the meeting, members will review the intercurrent literature to inform any updates. The units for update will be checklist items, research design categories, and LLM task categories delineated in the statement. At the meeting, the panel will discuss the current statement and suggest revisions considering public comments, literature review, and subject matter expertise. The steering committee will revise the statement based on this discussion, and this will be circulated to the expert panel for final review and approval. Review can result in the following action for each component of the TRIPOD-LLM statement (adapted from Mikati et al., 2019^33^) - items, research designs, and LLM tasks:

1. No modification
2. Modification of substantive content (small, editorial revisions such as re-wording for clarity and and correcting types will not be considered a modification)
3. Merging of one or more components together (merging will only take place within a component type)
4. Splitting one component into two or more components (splitting will only take place within a component type)
5. Retiring the component from the statement

Release of a new version of the statement will be disseminated to the community through postings on the TRIPOD-LLM website, the main TRIPOD website (https://www.tripod-statement.org/), the EQUATOR Network website (https://www.equator-network.org/reporting-guidelines/), and postings on social media accounts such as X (formerly known as Twitter) @TRIPODStatement. Emails will be sent to journal editors to inform them of the update and ensure that author instructions refer to the most current versioning. The detailed transcripts of the discussion are available in supplementary materials for full transparency.

At each review meeting, the membership of the expert panel will be reviewed for expertise, diversity, and representation, and new members will be solicited if and when gaps are identified. Expert panel members will also have the authority to trigger an ad hoc review of the guidelines to accommodate major, unexpected changes in the field that warrant more urgent discussion.

## Discussion

TRIPOD-LLM has been developed to guide researchers, journals, healthcare professionals, LLM developers (commercially and non-commercially), and healthcare institutions in the rapidly evolving field of biomedical and healthcare LLMs. It represents minimum reporting recommendations for studies describing LLMs’ development, tuning, or evaluation. Reporting TRIPOD-LLM items will help users understand and appraise the quality of LLM study methods, increase transparency around study findings, reduce overinterpretation of study findings, facilitate replication and reproducibility, and aid in implementing the LLM.

**Transparency throughout the model lifecycle** has been emphasized significantly in the guidelines. Detailed documentation is emphasized at each stage of an LLM’s life cycle;^37^ for example, during the development and fine-tuning phases, there is an emphasis on disclosing the origins and processing of training data. Moreover, the LLM version and specifics of any fine-tuning or alignment modeling processes on top of existing foundation models must be transparently reported to enable fair comparisons of LLMs. This includes specifying the cut-off dates for when training data were collected to clarify the temporal relevance of training datasets and potential for data leakage or contamination during evaluation. In addition, the model version date and if the model was frozen or remained dynamic during the data collection phase from generated output should be documented. Transparency regarding data is essential because LLMs are typically trained on multiple public large-scale datasets and thus inherently risk incorporating existing societal biases and inequities in stigmatizing language use as well as statistical risk allocation in disparate groups, necessitating a comprehensive transparency approach to training data sources and content to understand potential biases.^21,22,38–41^

**Human insight and oversight** are critical components of the TRIPOD-LLM statement, reflecting an emphasis on components eventually critical for the responsible deployment of LLMs (though deployment reliability and observability are outside the scope of this paper).^42–44^ The guidelines include requirements for increased reporting of the expected deployment context and specifying the levels of autonomy assigned to the LLM, if applicable. Furthermore, there is a focus on the quality control processes employed in dataset development and evaluation, such as qualifications of human assessors, requirement for dual annotation, and specific details on instructions provided to assessors to ensure that nuances of text evaluation are captured, thus facilitating reliable assessments of safety and performance.

**Prompting and task-specific performance** are key additions necessitated by the unique characteristics of LLMs. The variability in prompt engineering approaches can significantly influence LLM performance, potentially skewing benchmark comparisons and real-world applicability.^45,46^ Reports, where relevant, must include comprehensive descriptions of data sources used for developing prompts, LLM model name and versions, any preprocessing undertaken, and methods employed in prompt engineering. This ensures that prompts are effectively designed to elicit stable and reproducible performance from LLMs. Additionally, the guidelines call for clear reporting on evaluation settings, including instructions and interfaces used and characteristics of populations involved in evaluations. This is intended to ensure that LLM performance is assessed under conditions that closely mimic real-world applications, providing a reliable measure of its practical utility.

We anticipate that key users and beneficiaries of TRIPOD-LLM will be (1) academic and industry researchers authoring papers, (2) journal editors and peer reviewers evaluating research papers, and (3) other stakeholders (e.g., the research community in general, academic institutions, policy-makers, funders, regulators, patients, study participants, industry, and the broader public) who will benefit from increased transparency and quality of LLM research. We encourage editors, publishers, and the industry more broadly to support adherence to TRIPOD-LLM by referring to a link within the journal’s instructions to authors, enforcing its use during the submission and peer review process, and making adherence to the recommendations an expectation. We also encourage funders to require applications for LLM studies to include a plan to report their model according to the TRIPOD-LLM recommendations, thereby minimizing research waste and ensuring value for money.

Of note, this guideline was developed with text-only LLMs in mind; however, advances in multi-modal models incorporating LLMs, such as vision-language models,^47^ are now rapidly emerging - illustrating the need for rapid, nimble approaches for reporting guidelines. Many of the reporting considerations will be shared between text-only LLMs and these multi-modal models. For example, for vision-language models, both text and image preprocessing should be reported. However, unique considerations may arise that merit discussion in future versioning of TRIPOD-LLM or related guidelines. For example, studies reporting the development of LLMs that use imaging data should report details of image acquisition. In the interim, we suggest that studies reporting the development and/or evaluation of a method that includes an LLM as a primary component use the TRIPOD-LLM statement, although we acknowledge this may be subject to interpretation. We advise that users keep in mind the goals of reproducibility, understandability, and transparency to take a common-sense approach to deciding on the appropriate reporting guideline, and to interpreting the relevant components of TRIPOD-LLM statement items to report multimodal LLMs. Users may also refer to methodological guides from multiple AI fields, such as radiomics,^48,49^ to inform their reporting.

**The role of assurance labs** such as the Coalition for Health AI (CHAI)^50^ and Epic AI Labs,^51,52^ or internal validation standards are expected to be of importance in the generation, verification, certification, and maintenance of model cards for clinical AI. It is our opinion that the TRIPOD-LLM standard can and should inform such assurance labs as they develop approaches to assure LLMs in ways that meet the required regulatory bar for AI (e.g., the Biden-Harris Administration “Executive Order on Safe, Secure, and Trustworthy Artificial Intelligence”, the United States AI Safety Institute,^53^ the United States Office of the National Coordinator for Health Information Technology HTI-1 Final Rule,^54^ and the European Union AI Act^55^) and build confidence in patients, clinician, and other stakeholders about the utility and trustworthiness of clinical AI.

It must also be emphasized that LLM evaluation and validation requires specialized expertise and resources. To ensure equitable and safe deployment, investments into LLM development should be balanced by investments into infrastructure that enables robust validation beyond large academic settings. Moreover, this checklist should be seen as part of a continuous process for evaluating LLMs due to the temporal and geographic specific contexts these models inherit, which can impact the generalisability of performance and fairness across sites or at the same site over time. These shifts can be even more unpredictable than traditional ML models due to their user-dependent nature, and thus, significant effort must be placed on understanding trends and heterogeneity of effects instead of single-point estimates that proclaim universal validation.

### Conclusion

TRIPOD-LLM aims to assist authors in the complete reporting of their study and help LLM developers, researchers, peer reviewers, editors, policymakers, end-users (e.g., healthcare professionals), and patients understand data, methods, findings, and conclusions of LLM-driven research. Adhering to the TRIPOD-LLM reporting recommendations may promote the best and most efficient use of research time, effort, and money, enhancing the value of LLM research to maximize positive impact.

## Methods

The TRIPOD-LLM statement was formulated to guide the reporting of studies that develop, tune, or evaluate LLMs for any healthcare application or context and was crafted following pathways utilized in creating the other TRIPOD statements. An expedited Delphi process was implemented given the need for timely reporting guidelines in this field, and the living statement approach. An accompanying glossary (Box 1) defines essential terms relevant to the TRIPOD-LLM statement.

A steering group, including DSB, JG, LAC, GSC, and KGMM, was established to direct the guideline development process. They were joined by an expert panel, including SC, CF, DR, GS, TM, DFD, RU, LHH, YA, JWG, LGM, NM, and RD, and were chosen based on their diverse expertise and experience in natural language processing, artificial intelligence, and medical informatics. This guideline was registered on May 2, 2024, with the EQUATOR Network as a reporting guideline under development (www.equator-network.org).

### Ethics

This study received an exemption from the MIT COUHES IRB review board on March 26, 2024 (Exempt ID: E-5705). Delphi survey participants provided electronic informed consent before completing the survey.

### Candidate item list generation

The TRIPOD-2015 and TRIPOD+AI guidelines (www.tripod-statement.org) and associated literature on reporting guidelines for LLMs were used to inform the initial candidate item list drafted by DSB and JG.^9,11,28,32^ The steering group and expert panel expanded this list through additional literature review, ultimately standardizing it to 64 unique items across the following sections: title, abstract, introduction, methods, results, discussion, and others.

### Panelist recruitment

Delphi participants were identified by the steering committee from authors of relevant publications and through personal recommendations, including experts recommended by other Delphi participants. The steering group identified participants representing geographical and disciplinary diversity, including key stakeholder groups, e.g., researchers (statisticians, data scientists, epidemiologists, machine learners, clinicians, and ethicists), healthcare professionals, journal editors, funders, policymakers, healthcare regulators, and patient advocate groups. No minimum sample size was placed on the number of participants. A steering group member checked the expertise or experience of each identified person. Participants were then invited to complete a survey via e-mail. Delphi participants did not receive any financial incentive or gift to participate.

### Delphi process

The survey was designed to allow individual responses, in English and delivered electronically using Google Forms. All responses were anonymous; no emails or identifying information was collected from participants. Participants were asked to rate each item as ‘can be omitted,’ ‘possibly include,’ ‘desirable for inclusion,’ or ‘essential for inclusion’ as has been conducted in previous TRIPOD guidelines.^9^ Participants were also invited to comment on any item and suggest new items. DSB and JG collated and analyzed the free text responses then used the themes generated to inform item rephrasing, merging, or suggesting new items. All steering group members were invited to participate in the Delphi surveys.

### Round 1 participants

The first round was conducted between 01 March 2024 and 23 April 2024, where the participation link was sent to 56 people. Of the 56 invited, 26 completed the survey. Survey participants came from 9 countries, of which 14 were from North America, five from Europe, two from Asia, one from South America, and one from Australasia. Three participants did not provide this information. Participants reported their primary fields of work and could select more than one field. 20/26 (77 %) had a primary field in AI, Machine Learning, Clinical Informatics, or NLP, and 14/26 selected healthcare.

### Consensus Meeting

An online consensus meeting was held on the 22nd and 24th of April, chaired by DSB and JG via Zoom. All steering committee and expert panel members were invited to attend. Recordings and notes were sent immediately after the meetings to enable asynchronous contribution for those who did not attend. The responses to each question were examined in turn, as well as all free-text comments. Items with <50% ‘Essential to Include’ were highlighted and deliberated for the importance of inclusion. Agreement by consensus without needing a third party was reached in all cases. To arrive at a consensus, the item was discussed until no panel had additional comments or disagreements with the final disposition of the item.

Due to the vast number of applications being developed using LLMs, a modular approach was used to group included items under additional subcategories under the ‘Research Design’ and ‘LLM Task’ headers. This approach was agreed upon during the meeting, and the steering committee approved the final groupings.

To adequately address the variety of studies and uses regarding LLMs, ranging across stages of development, tuning, evaluation, and implementation, as well as across healthcare tasks, items are categorized according to (1) research design and (2) LLM task. The research design categories are: *de novo* LLM development, LLM methods such as fine-tuning, prompt-engineering techniques and architecture modifications, intrinsic LLM evaluation, and LLM evaluation in dedicated healthcare settings and tasks. The LLM task categories are lower-level text processing (e.g., part-of-speech tagging, relation extraction, named entity recognition), classification (e.g., diagnosis), long-form question-answering, conversational agent, documentation generation, summarization/simplification, machine translation, and outcome forecasting (e.g., prognosis). Items may apply to several design and task categories, and studies may include more than one design and task. Items applicable to every type and task covered in the study should be reported. Definitions and examples of each design and task category are provided in Table 1. We acknowledge that these categorizations are imperfect and overlap may exist across designs and tasks.

## Data Availability

All data produced in the present work are contained in the manuscript.

https://tripod-llm.vercel.app/

## Acknowledgments

JG is funded by the NIH-USA U54 TW012043-01 and NIH-USA OT2OD032701.

SC was supported by NIH-USA U54CA274516-01A1.

GSC was supported by Cancer Research UK (programme grant: C49297/A27294), and by EPSRC grant for ‘Artificial intelligence innovation to accelerate health research’ (number: EP/Y018516/1, and is a National Institute for Health and Care Research (NIHR) Senior Investigator. The views expressed in this article are those of the author and not necessarily those of the NIHR, or the Department of Health and Social Care.

Yin Aphinyanaphongs was partially supported by NIH 3UL1TR001445-05 and National Science Foundation award #1928614 & #2129076.

LAC is funded by NIH-USA U54 TW012043-01, NIH-USA OT2OD032701, and NIH-USA R01EB017205.

DSB was supported by NIH-USA U54CA274516-01A1 and NIH-USA R01CA294033-01.

Dr. Gichoya is a 2022 Robert Wood Johnson Foundation Harold Amos Medical Faculty Development Program and declares support from RSNA Health Disparities grant (#EIHD2204), Lacuna Fund (#67), Gordon and Betty Moore Foundation, NIH (NIBIB) MIDRC grant under contracts 75N92020C00008 and 75N92020C00021, and NHLBI Award Number R01HL167811.

