## Supplemental Table 3 for "The TRIPOD-LLM Statement: A Targeted Guideline For Reporting Large Language Models Use"

**Supplementary Table 3:** TRIPOD-LLM Expanded Checklist (Explanation and Elaboration Light)

| **Section** | **Item** | **Checklist Item** | **Research Design** | **LLM Task** |
| --- | --- | --- | --- | --- |
| Title | | | | |
| Title | 1 | **Identify the study as developing, fine-tuning, and/or evaluating the performance of an LLM, specifying the task, the target population, and the outcome to be predicted.**   - Informative titles help with the identification of LLM-based studies by potential readers and also systematic reviewers - Report an informative title that provides important information about the target population and the outcome | All | All |
| Abstract |  |  |  |  |
| Abstract | 2 | **See TRIPOD-LLM Abstract**   - Report an abstract addressing each item in the TRIPOD-LLM for Abstracts checklist | All | All |
| Introduction |  |  |  |  |
| Background | 3a | **Explain the healthcare context / use case (e.g., administrative, diagnostic, therapeutic, clinical workflow) and rationale for developing or evaluating the LLM, including references to existing approaches and models.**   - Describe the healthcare setting or use case where the LLM is intended to be used. - Where existing approaches or LLMs are available, provide a clear justification for developing a new LLM. - For studies evaluating an existing model, provide the rationale for the evaluation and references to all models being evaluated. - For *de novo* LLM development and LLM methods studies, the precise healthcare context/use cases may not be determined. In this case, provide examples of potential future healthcare contexts/use cases. | All | All |
|  | 3b | **Describe the target population and the intended use of the LLM in the context of the care pathway, including its intended users in current gold standard practices (e.g., healthcare professionals, patients, public, or administrators).**   - Describe who the target population is for the developed or evaluated LLM, such as people of a certain age, in a specific country, or with a specific disease. - Describe the intended purpose of the LLM, including the clinical decision or guidance the LLM is intended to support (e.g., referral for further testing or hospital admission, triage, starting a treatment, patient portal messaging, billing) and the point in the care pathway where the LLM is intended to be used. - Describe who the intended users of the LLM are, and whether the LLM is for healthcare professionals, patients, public, or other stakeholders. - Explain the current gold standard practices that this LLM is seeking to interact with or replace. | E  H | All |
| Objectives | 4 | **Specify the study objectives, including whether the study describes the initial development, fine-tuning, or validation of an LLM (or multiple stages).**   - Provide an explicit statement of all objectives of the study, describing whether the study is developing an LLM, fine-tuning or otherwise adjusting an existing LLM, incorporating an existing LLM within a new informatics pipeline or framework, evaluating the performance of an LLM, or covering multiple stages. | All | All |
| **Methods** | | | | |
| Data | 5a | **Describe the sources of data separately for the training, tuning, and/or evaluation datasets and the rationale for using these data (e.g., web corpora, clinical research/trial data, EHR data).**   - Provide transparency about the data sources used, including whether the data are, for example, from specific web sources, a randomized trial, a registry or from electronic routine healthcare records - Specify whether the study is using existing data or is prospectively collecting new data for the purpose of LLM updating, finetuning or evaluation - Where existing data are being used (i.e., they were originally collected for a different purpose), provide the rationale for using these data, and comment on the suitability (particularly if data are being used from a different setting, country, and/or clinical population to the intended target population) and representativeness of these data with respect to the intended target population and context - If any synthetic data have been used, then provide reasons as to why, and provide all details on how the synthetic data have been created (and code, see item 14f) and used in the study | All | All |
|  | 5b | **Describe the relevant data points and provide a quantitative and qualitative description of their distribution and other relevant descriptors of the dataset (e.g., source, languages, countries of origin)**   - Offer a comprehensive understanding of the dataset used, relevant metadata, languages, and breakdown of characteristics. - Include both quantitative and qualitative descriptions of the data. | All | All |
|  | 5c | **Specifically state the date of the oldest and newest item of text used in the development process (training, fine-tuning, reward modeling) and in the evaluation datasets.**   - Ensure the temporal relevance and validity of the data used for training and/or evaluation. - Provide dates for the text items used in different stages of development and evaluation. - For studies using existing LLMs, provide reference(s) to this information if provided by the original developers or state that this information is not available. | All | All |
|  | 5d | **Describe any data pre-processing and quality checking, including whether this was similar across text corpora, institutions, and relevant socio-demographic groups.**   - If data cleaning is performed e.g. from raw EHR notes, describe any data cleaning steps. This includes transformations of raw data, data quality checks, or translation. All code used for data cleaning should be made available (see item 14e). - Report any efforts in mitigating biased or false content in training. - Report feature selection techniques, if any. - If the data pre-processing/data cleaning steps are extensive, consider reporting this information in the supplementary material. | All | All |
|  | 5e | **Describe how missing and imbalanced data were handled and provide reasons for omitting any data.**   - If the data used are linked with other data or have the potential for missingness (e.g., when extracted from EHR), report any missingness overall and across groups. - If individuals’ data have been omitted due to missing values, this should be reported, and reasons given. Note that this is generally not applicable for LLM pretraining. | All | All |
| Analytical Methods | 6a | **Report the LLM name, version, and last date of training.**   - Given the rapid pace of the field, clear details about the type and version of model used aid in fair comparison across different studies. - For studies using existing LLMs, provide reference(s) to this information if provided by the original developers or state that this information is not available. | All | All |
|  | 6b | **Report details of LLM development process, such as LLM architecture, training, fine-tuning procedures, and alignment strategy (e.g., reinforcement learning, direct preference optimization, etc.) and alignment goals (e.g., helpfulness, honesty, harmlessness, etc.).**   - Outline the complete development process and alignment strategies that were implemented in this study, or point to a study that describes this process. - For any fine-tuning approach, provide details on hyperparameter search and settings, and type of fine-tuning (e.g. full fine-tuning, parameter-efficient fine-tuning strategies). - Specify alignment goals for the LLM and what instructions were given to any labelers involved with the alignment process. | M  D | All |
|  | 6c | **Report details of how text was generated using the LLM, including any prompt engineering and inference settings (e.g., seed, temperature, max token length, penalties), as relevant.**   - Describe the model architecture and configuration. - Include details on inference settings such as parameters that control generation, including how these settings were arrived at (e.g., type of sampling used, beam-search). - Provide details on any use of constrained decoding, and any post-processing applied to generated text. | M  D  E | All |
|  | 6d | **Specify the initial and post-processed output of the LLM (e.g., probabilities, classification, unstructured text).**   - Specify whether the outputs are probabilities, classifications, or unstructured text. - Explain how the initial outputs are transformed or refined in the post-processing stage. All code used for post-processing should be made available (see item 14e). - If the post-processing steps are extensive, consider reporting this information in the supplementary material. | All | All |
|  | 6e | **Provide details and rationale for any classification and, if applicable, how the probabilities were determined and thresholds identified.**   - Describe the process and criteria for categorizing outputs into different classes or groups. - Specify the algorithms or formulas used to derive probability estimates. - Provide a rationale for the chosen thresholds, referencing literature, clinical guidelines, statistical considerations, or ad-hoc decisions. | All | C  OF |
| LLM Output | 7a | **Include metrics that capture the quality of generative outputs, such as consistency, relevance, similarity, and accuracy, compared to gold standards.**   - Given the stochastic nature of LLMs, metrics like consistency, relevance, similarity, and accuracy aid in providing improved characterisation of the results. - Explain how the generative outputs are measured against established benchmarks or reference standards. - Define what gold standard was used or what algorithms or scores derived such metrics. - Provide details of how consistency is measured, e.g. variability to different prompt variations. | All | QA  IR  DG  SS  MT |
|  | 7b | **Report the outcome metrics' relevance to downstream task at deployment time and correlation of metric to human evaluation of the text for the intended use.**   - Describe how the outcome metrics are relevant to the real-world application of the LLM. - If human evaluation is carried out, explain how these metrics correlate with human assessments of the text, ensuring the outputs meet user expectations and requirements. | E  H | All |
|  | 7c | **Clearly define the outcome, how the LLM predictions were calculated (e.g., formula, code, object, API), the date of inference for closed-source LLMs, and evaluation metrics.**   - Describe the methodology used for generating LLM output. Include details such as whether a specific algorithm, codebase, software object, or API was used. - Closed-source LLMs may be updated without changes in the named versioning. To enable fair comparisons, report the date of inference for closed-source LLMs. | E  H | All |
|  | 7d | **If outcome assessment requires subjective interpretation, describe the qualifications of the assessors, any instructions provided, relevant information on demographics of the assessors, and inter-assessor agreement.**   - Provide information on the assessors’ professional background and expertise relevant to the task. - Describe the guidelines and criteria provided to the assessors for the evaluation process. - Include information about the assessors’ demographics to ensure diversity and representativeness. - Report the level of agreement among the assessors using appropriate statistical measures. | All | All |
|  | 7e | **Specify how performance was compared to other LLMs, humans, and other benchmarks or standards.**   - Explain the process and criteria for comparing the LLM’s performance with other models and how these are and are not fair comparisons. - Detail how LLM performance was compared to humans and any differences in the generation and evaluation process between the two groups | All | All |
| Annotation | 8a | **If annotation was done, report how text was labeled, including providing specific annotation guidelines with examples.**   - Provide a copy of the annotation guidelines provided to any annotators along with any examples. - Provide any other training or reference material provided to the annotators. | All | All |
|  | 8b | **If annotation was done, report how many annotators labeled the dataset(s), including the proportion of data in each dataset that were annotated by more than 1 annotator, and the inter-annotator agreement.**   - State the number of annotators that were used in total, and what proportion was annotated by multiple individuals - Report the level of agreement among annotators when multiple were used using appropriate statistical measures e.g. Cohen’s kappa. | All | All |
|  | 8c | **If annotation was done, provide information on the background and experience of the annotators.**   - Provide details on the professional background, qualifications, and experience of the annotators. | All | All |
| Prompting | 9a | **If research involved prompting LLMs, provide details on the processes used during prompt design, curation, and selection.**   - Describe the methodology used to create the initial set of prompts. - Explain the criteria and process used to refine and curate the prompts. - Detail the process used to select the final set of prompts from the curated list. - Describe how prompts were tested to ensure they effectively elicited the desired responses from the LLM. | All | All |
|  | 9b | **If research involved prompting LLMs, report what data were used to develop the prompts.**   - Describe the datasets or sources of information used to create the prompts. - Provide details on the datasets used to evaluate the performance of the prompts. - Report if there was any overlap between the datasets used to develop the prompts and to evaluate the methods. | All | All |
| Summarization | 10 | **Describe any preprocessing of the data before summarization.**   - Outline any preprocessing steps applied to the data before summarization e.g. de-identification. - State if any reformatting or additional processing was performed specifically for summarization e.g. removal of specific sections. | All | SS |
| Instruction tuning/Alignment | 11 | **If instruction tuning/alignment strategies were used, what were the instructions and interface used for evaluation, and what were the characteristics of the populations doing evaluation?**   - Describe the specific instruction/preference datasets provided to the LLM during the tuning or alignment process. - Describe the interface or tools through which evaluators evaluate and provide feedback on the LLM's performance during alignment. - Provide information on the demographics and expertise of the evaluators. | M  D | All |
| Compute | 12 | **Report compute, or proxies thereof (e.g., time on what and how many machines, cost on what and how many machines, inference time, floating-point operations per second (FLOPs)), required to carry out methods.**   - Specify the computational resources used, for example machines, time, and cost. - Report the inference time and any metrics related to computational efficiency, as available. - If possible, provide additional metrics such as FLOPs to quantify the computational requirements. | M  D  E | All |
| Ethical Approval | 13 | **Name the institutional research board or ethics committee that approved the study and describe the participant-informed consent or the ethics committee waiver of informed consent.**   - Name the institutional review board or ethics committee that provided approval. - Describe the informed consent process or the waiver granted by the ethics committee. | All | All |
| Open Science | 14a | **Give the source of funding and the role of the funders for the present study.**   - Identify the funding sources supporting the research. - Describe any role the funders had in the study design, data collection, analysis, or publication. | All | All |
|  | 14b | **Declare any conflicts of interest and financial disclosures for all authors.**   - Disclose any relationships or activities that could be perceived as influencing the research. - Provide information on any financial interests or affiliations of the authors. | All | All |
|  | 14c | **Indicate where the study protocol can be accessed or state that a protocol was not prepared.**   - Provide details on where and how the clinical study protocol can be accessed by others. | H | All |
|  | 14d | **Provide registration information for the study, including register name and registration number, or state that the study was not registered.**   - If a clinical trial component is undertaken, state the name of the registry and the registration number for the study. - Clearly state if the study was not registered and provide reasons if applicable. | H | All |
|  | 14e | **Provide details of the availability of the study data.**   - Explain where and how the study data can be accessed, including any conditions or restrictions. | All | All |
|  | 14f | **Provide details of the availability of the code to reproduce the study results.**   - Describe how and where the code used in the study can be accessed by others. | All | All |
| Public Involvement | 15 | **Provide details of any patient and public involvement during the design, conduct, reporting, interpretation, or dissemination of the study or state no involvement.**   - Describe how patients or the public were involved in various stages of the research. - Explain if and how the findings were shared with patients or the public. | H | All |
| **Results** | | | | |
| Participants | 16a | **When using patient/EHR data, describe the flow of text/EHR/patient data through the study, including the number of documents/questions/participants with and without the outcome/label and follow-up time as applicable.**   - If EHR data is used, describe the process of how patient/EHR data were selected, filtered, and included in the study. - Specify the number of documents, questions, or participants included and excluded at each stage. - Indicate the number of participants with and without the specific outcome/label and the duration of follow-up. | E  H | All |
|  | 16b | **When using patient/EHR data, report the characteristics overall and, for each data source or setting, and for development/evaluation splits, including the key dates, key characteristics, and sample size.**   - Provide a summary of the overall demographic and clinical characteristics of the dataset. - Detail the characteristics for each specific data source or setting. - Describe the sample size and key characteristics for the development and evaluation datasets. | E  H | All |
|  | 16c | **For LLM evaluation that include clinical outcomes, show a comparison of the distribution of important clinical variables that may be associated with the outcome between development and evaluation data, if available.**   - Provide a comparison of key predictors, demographics, and clinical characteristics between the development and evaluation datasets. - Report whether the distribution of predictors, demographics, and clinical is comparable between datasets. - These characteristics will depend on the specific context of use and task for each study, as established by literature review and/or domain expert input. | E  H | All |
|  | 16d | **When using patient/EHR data, specify the number of participants and outcome events in each analysis (e.g., for LLM development, hyperparameter tuning, LLM evaluation).**   - Report the number of participants and outcome events for each specific analysis. - Describe the stages of analysis and the corresponding data used. | E  H | All |
| Performance | 17 | **Report LLM performance according to pre-specified metrics (see item 7a) and/or human evaluation (see item 7d).**   - Report performance overall and for any key subgroups (e.g., sociodemographic, diagnosis, data source). - Consider plots to aid presentation. - Consider reporting confidence intervals overall and for any key subgroups. - Consider reporting uncertainty estimation of the generated output (e.g., LLM-verbalized estimates, logit-based estimates) overall and for any key subgroups. | All | All |
| LLM Updating | 18 | **If applicable, report the results from any LLM updating, including the updated LLM and subsequent performance.**   - Explain any modifications or updates made to the LLM and the reasons behind them. - Report the performance metrics of the updated LLM. | All | All |
| **Discussion** | | | | |
| Interpretation | 19a | **Give an overall interpretation of the main results, including issues of fairness in the context of the objectives and previous studies.**   - Summarize the main findings and their implications overall and for the specified or anticipated healthcare contexts of use. - Discuss any fairness or robustness issues observed, such as biases in predictions. | All | All |
| Limitations | 19b | **Discuss any limitations of the study and their effects on any biases, statistical uncertainty, and generalizability.**   - Identify and explain the limitations of the study design, robustness of results, and implications for generalisability of findings. | All | All |
| Usability of the LLM in context | 19c | **Describe any known challenges in using data for the specified task and domain context with reference to representation, missingness, harmonization, and bias.**   - Explain the difficulties encountered in using the data for the specified task e.g. formatting inconsistencies, missingness, class imbalance, or harmonization challenges. - Discuss issues related to data representation and potential biases that may impact findings generalizability or robustness. | E  H | All |
|  | 19d | **Define the intended use for the implementation under evaluation, including the intended input, end-user, level of autonomy/human oversight.**   - Specify the purpose of the LLM and the type of input it requires. - Describe the end-users and the level of autonomy or human oversight required. - Discuss barriers to access by the intended end-user, e.g. lack of access to hospital systems with EHRs, wifi, technical support. | E  H | All |
|  | 19e | **If applicable, describe how poor quality or unavailable input data should be assessed and handled when implementing the LLM, i.e., what is the usability of the LLM in the context of current clinical care.**   - Explain strategies for managing poor quality or missing input data. - Describe the LLM’s usability in real-world clinical settings. | E  H | All |
|  | 19f | **If applicable, specify whether users will be required to interact in the handling of the input data or use of the LLM, and what level of expertise is required of users.**   - Describe the extent of user interaction needed for handling input data or operating the LLM. - Specify the level of expertise needed to use the LLM effectively. | E  H | All |
|  | 19g | **Discuss any next steps for future research, with a specific view to applicability and generalizability of the LLM.**   - Outline potential areas for further investigation to improve the LLM. - Discuss how the findings can be applied to other contexts or populations | All | All |

### LLM = large language model; M = LLM methods; D = *de novo* LLM development; E = LLM evaluation; H = LLM evaluation in healthcare settings; C = classification; OF = outcome forecasting; QA = long-form question-answering; IR = information retrieval; DG = document generation; SS = summarization and simplification; MT = machine translation, EHR = electronic health record.
